# Meta-analysis of gestational duration and spontaneous preterm birth identifies new maternal risk loci

**DOI:** 10.1101/2022.10.31.22281753

**Authors:** A. Pasanen, M. K. Karjalainen, FinnGen, G. Zhang, H. Tiensuu, A. M. Haapalainen, M. Ojaniemi, B. Feenstra, B. Jacobsson, A. Palotie, H. Laivuori, L. J. Muglia, M. Rämet, M. Hallman

## Abstract

**Background:** Preterm birth (<37 weeks of gestation) is a major cause of neonatal death and morbidity. Up to 40% of the variation in timing of birth results from genetic factors, mostly due to the maternal genome.

**Methods:** We conducted a genome-wide meta-analysis of gestational duration and spontaneous preterm birth in 68,732 and 98,371 European mothers, respectively.

**Results:** We detected 19 associated loci of which seven were novel. The loci mapped to several biologically plausible genes, including *HAND2* whose expression was previously shown to decrease during gestation, associated with gestational duration, and *GC* encoding Vitamin D-binding protein, associated with preterm birth. Downstream *in silico*-analysis suggested regulatory roles as underlying mechanisms for the associated loci. LD score regression found birth weight measures as the most strongly correlated traits, highlighting the unique nature of spontaneous preterm birth phenotype. Tissue expression and colocalization analysis revealed reproductive tissues and immune cell types as the most relevant sites of action.

**Conclusion:** We report novel genetic risk loci that associate with preterm birth or gestational duration, and reproduce findings from previous genome-wide association studies. Altogether, our findings provide new insight into the genetic background of preterm birth. Better characterization of the causal genetic mechanisms will be important to public health as it could suggest new strategies to treat and prevent preterm birth.

---

Proper timing of birth is crucial for the survival and long-term health of newborn infants. Preterm birth, defined as birth that occurs prior to 37 completed weeks of gestation, is the most common cause of neonatal death and a prevalent cause of death among children under 5 years^1^. Moreover, preterm birth is the underlying cause of several long-term morbidities including neurodevelopmental problems, cerebral palsy, learning difficulties, and sensory loss^2^. Globally, preterm birth affects approximately 11% of births, equal to 15 million pregnancies, each year. In Scandinavian countries and Finland, the annual incidence of preterm birth is approximately 5–6%^2,3^.

While intrauterine growth restriction and preeclampsia are major causes of medically indicated preterm birth, approximately 70% preterm births occur after spontaneous onset of labor^1^. There are just a few ways to predict the risk or to prevent the occurrence of spontaneous preterm birth (SPTB)^4^. Genetic variants in maternal and fetal genomes have been recognized as factors that contribute to the risk of SPTB and to variation in gestational duration. Family studies suggest that approximately 30%–40% of the variation in timing of birth is explained by genetic factors, with contributions from the maternal genome most important^5–7^. Recent genome-wide association studies (GWAS) have identified some robust associations. Variants in genes including *WNT4, EBF1, AGTR2*, and *KCNAB1* were associated with timing of birth in mothers, and a fetal GWAS meta-analysis discovered a locus near genes that encode pro-inflammatory cytokines associated with gestational duration^8–10^.

In the present study, our aim was to strengthen knowledge of the genetic background of SPTB by identifying and replicating associations of genetic loci in relation to timing of spontaneous singleton birth. To that end, we conducted a case-control meta-analysis of SPTB and a quantitative meta-analysis of gestational duration in 98,371 and 68,732 European mothers, respectively.

## Results

### Overview of genome-wide meta-analysis

The genome-wide meta-analysis of SPTB (n=98,371) and gestational duration (n=68,732) with maternal data from the FinnGen study, 23andMe, Inc., and the cohort from Northern and Central Finland detected 19 independent loci with at least one variant associated at *p*<5e−8 (Figure 1a, Tables 1-2). Variants associated with gestational duration were enriched in several categories, whereas variants associated with SPTB were mostly annotated as intronic or intergenic (Figure 1b). We considered an associated locus to be novel if there were no genome-wide significant associations with gestational duration or SPTB for any of the variants within a ±1 Mb range around the meta-analysis lead variant in the GWAS Catalog^11^ (Table S2) or in a recent maternal meta-analysis of the timing of parturition^10^. We detected five novel loci associated with gestational duration, and two novel risk loci for SPTB. The results of the meta-analysis with a strict SPTB definition in the FinnGen GWAS are shown in Figure S1. The effect estimates of the associated loci were similar with the main analysis (Figure S2).

**Figure 1.**
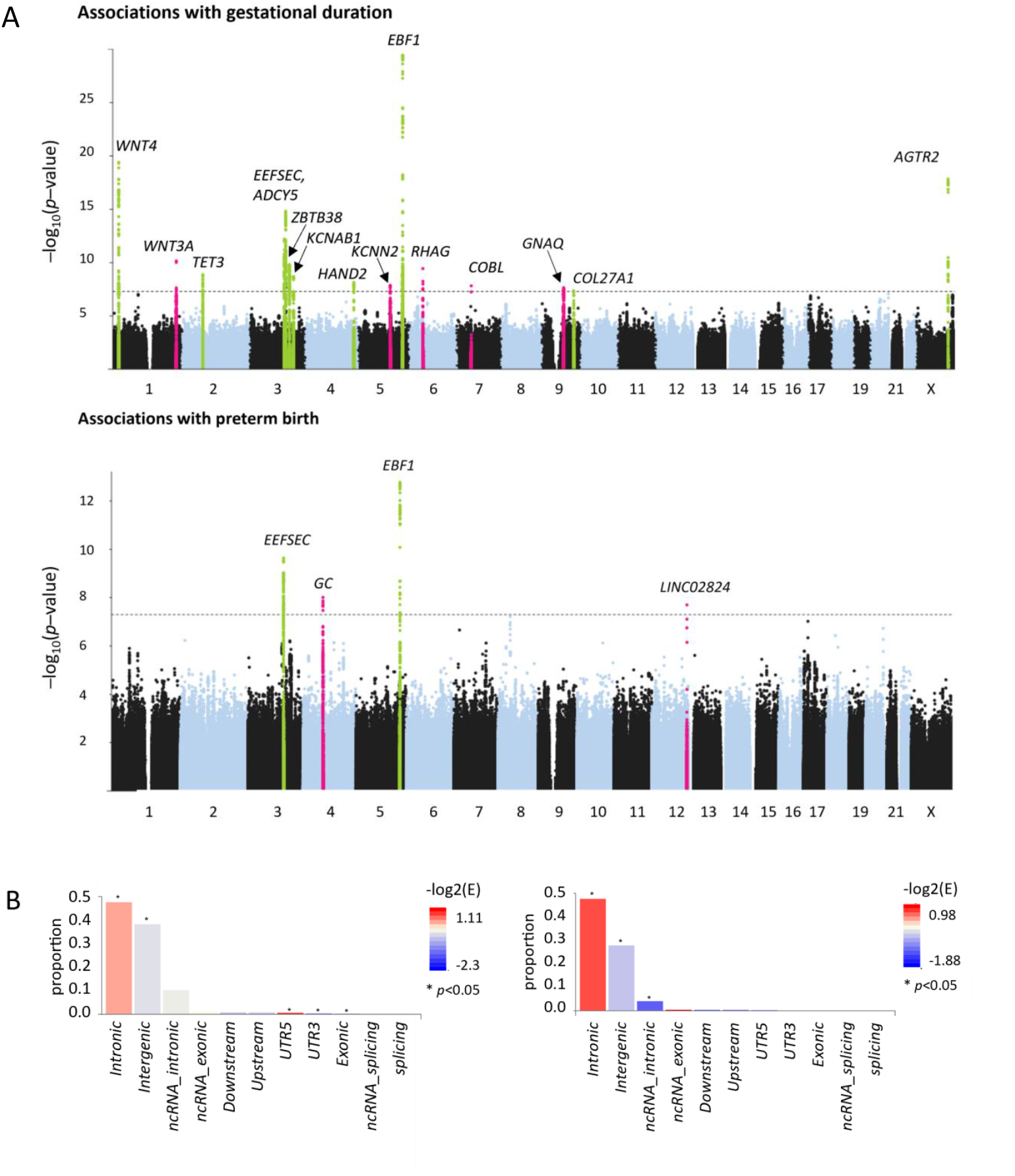
Meta-analysis of gestational duration and SPTB. **A)** Loci with genome-wide significant associations (*p* < 5e-08) are highlighted in the Manhattan plots. Chromosomal positions are shown at the x-axis, and the y-axis shows association *p* values at the -log_10_ scale. The meta-analysis detected 14 loci associated with gestational duration and four loci associated with SPTB. Peaks highlighted in pink represent novel loci, and peaks highlighted in green show known loci. **B)** Annotations as number of SNPs per functional consequences on genes.

**Table 1.** Loci associated with gestational duration in meta-analysis of 68,732 women of European ancestry. Loci highlighted in bold had no previous associations (*p* < 5e-8) with gestational duration or preterm birth.

| Chr:Pos | Rsid | A1 | Freq | Z-score | p-value | HetPVal | Gene |
| --- | --- | --- | --- | --- | --- | --- | --- |
| **5:158460958** | rs6881817 | T | 0.244 | -11.41 | 3.74E-30 | 0.726 | *EBF1* |
| **1:22141722** | rs3820282 | T | 0.149 | 9.189 | 3.95E-20 | 0.075 | *WNT4* |
| **23:116053119** | rs5950512 | A | 0.401 | -8.791 | 1.48E-18 | 0.512 | *AGTR2* |
| **3:128159974** | rs2999049 | T | 0.720 | -7.972 | 1.56E-15 | 0.258 | *EEFSEC* |
| **3:123365694** | rs10934646 | A | 0.634 | -7.193 | 6.35E-13 | 0.121 | *ADCY5* |
| **1:228015567** | **rs708119** | **C** | **0.670** | **6.514** | **7.32E-11** | **0.207** | ***WNT3A*** |
| **3:141414608** | rs1991431 | A | 0.441 | 6.402 | 1.54E-10 | 0.583 | *ZBTB38* |
| **6:49592080** | **rs10948514** | **T** | **0.274** | **6.267** | **3.68E-10** | **0.999** | ***RHAG*** |
| **2:74009288** | rs71848031 | D | 0.057 | -6.061 | 1.36E-09 | 0.913 | *TET3* |
| **3:156137629** | rs4679760 | C | 0.426 | -5.992 | 2.07E-09 | 0.360 | *KCNAB1* |
| **4:173813320** | rs7697038 | A | 0.323 | 5.780 | 7.45E-09 | 0.355 | *HAND2* |
| **5:114208039** | **rs13175113** | **T** | **0.486** | **5.668** | **1.45E-08** | **0.174** | ***KCNN2*** |
| **7:51940143** | **rs151143987** | **T** | **0.996** | **5.655** | **1.56E-08** | **0.712** | ***COBL*** |
| **9:77936015** | **rs11145617** | **A** | **0.210** | **-5.586** | **2.32E-08** | **0.584** | ***GNAQ*** |
| **9:114160401** | rs2808791 | T | 0.474 | 5.467 | 4.58E-08 | 0.429 | *COL27A1* |

Chr, chromosome; Pos, position (Grch38); A1, effect allele; HetPval, $p$ value for heterogeneity. Novel loci are highlighted in bold; reported genes are the nearest genes. For replicated loci, candidate genes are based on previous studies.

**Table 2.**
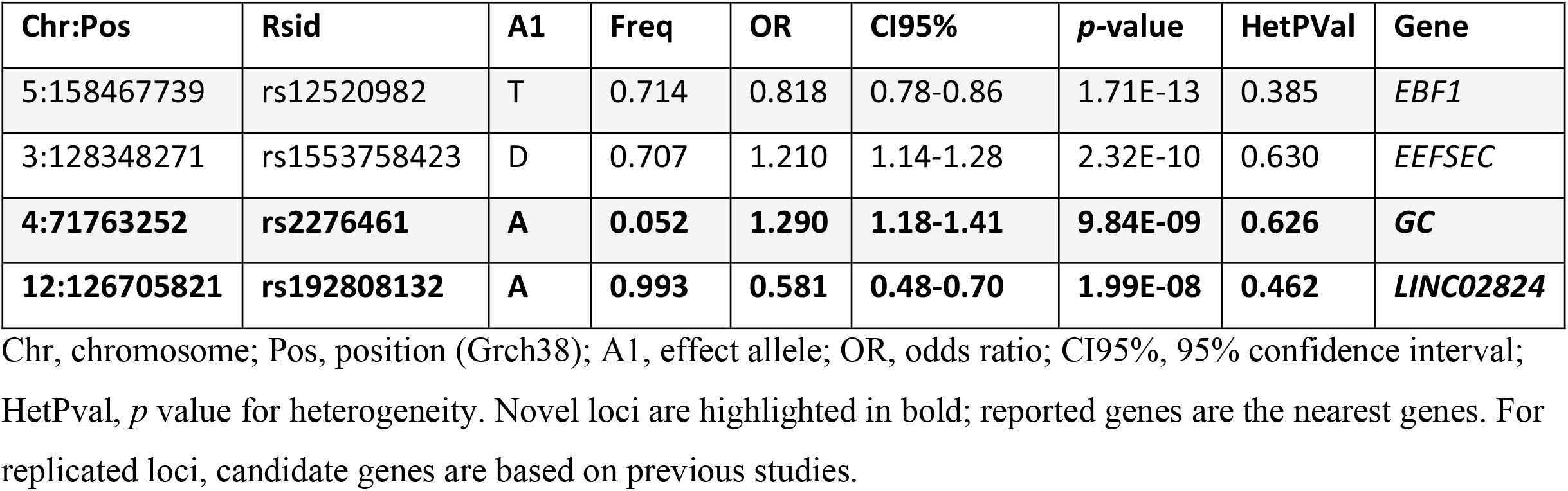
Loci associated with SPTB in meta-analysis of 98,372 women of European ancestry. Loci highlighted in bold had no previous associations (*p* < 5e-8) with gestational duration or preterm birth.

LDSC-based genomic inflation factor^12^ indicated minimal confounding effects in the meta-analysis of gestational duration (λ_GC_=1.077, intercept=1.025) or SPTB (λ_GC_=1.038, intercept=1.007) (Figure S3). The meta-analysis test statistics were homogenous among populations, and the effect estimates of the risk loci for SPTB were similar across individual cohorts (Tables 1-2, Figure S4). According to LDSC heritability estimates, the current results explain approximately 17.5% of the variation in gestational duration and 6% in SPTB on a liability scale (Table S3). We further used LDSC to evaluate shared genetic architecture between the meta-analysis outcomes and 773 other complex traits (Figure 2, Table S4). The analysis detected significant correlations between birth weight–related measures and both gestational duration and SPTB. As expected, longer duration of gestation was associated with higher birth weight, whereas preterm birth was linked to lower birth-weight measures. In addition, specific measures of physical fitness, alertness, and lack of depression correlated with a longer duration of pregnancy or term birth (Figure2, Table S4).

**Figure 2.**
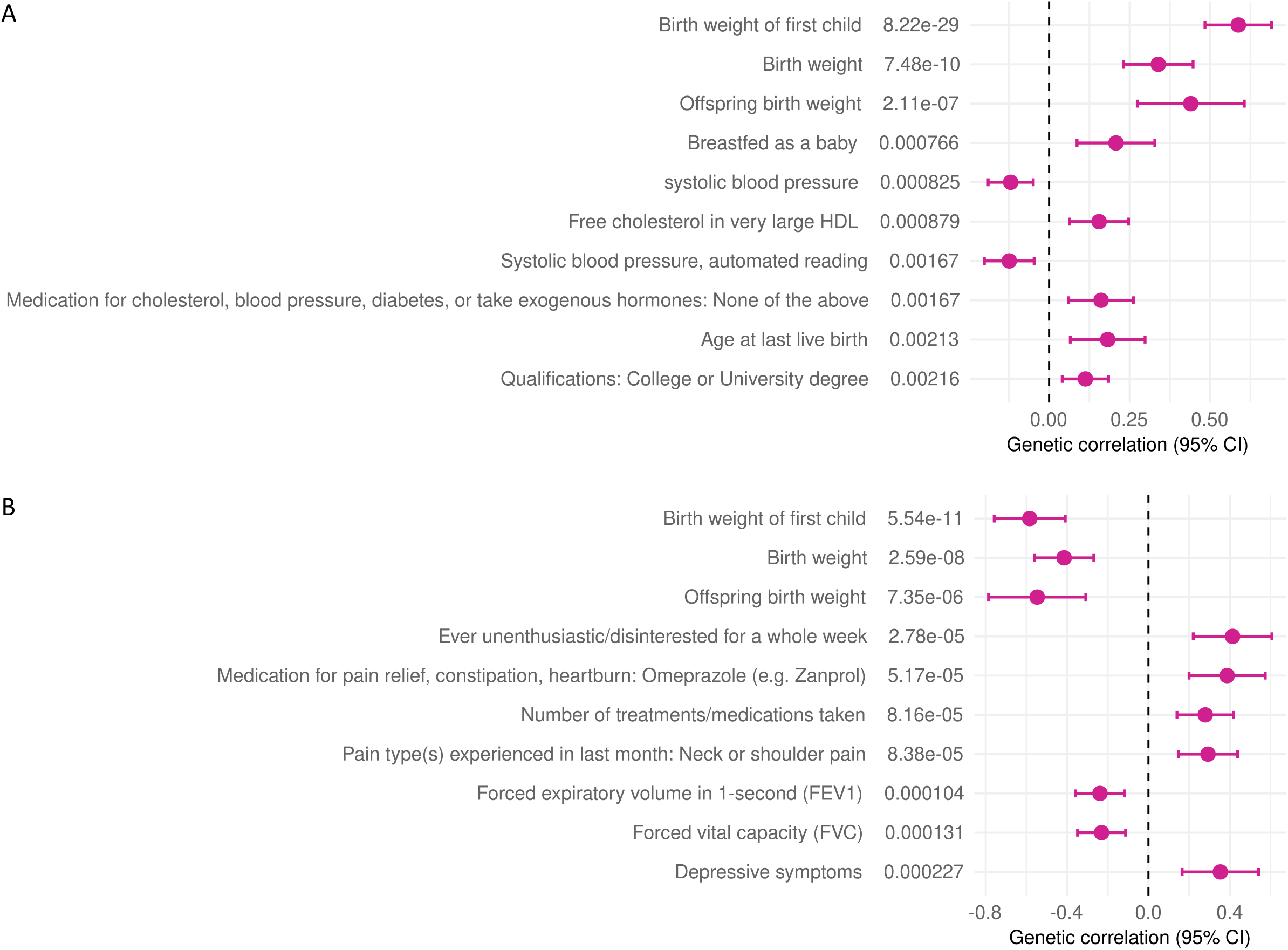
Genetic correlations between A) gestational duration or B) SPTB and other complex traits. Genetic correlation between gestational duration or SPTB and a comprehensive set of 773 complex traits was analyzed with LD score regression. Top 10 correlated traits, followed by their respective *p* values, are shown.

MAGMA gene set enrichment analysis based on the full distribution *p-*values indicated involvement of gonad development and steroid hormone biosynthesis in gestational duration, whereas kinetochore-microtubule and neuron differentiation were the top pathways in SPTB (Table S5). MAGMA tissue expression analysis across GTEx v8 did not yield significant results but ranked several reproductive tissues, including uterus and ovary, among the most relevant tissue types for both gestational duration- and SPTB-associated genes (Figure S5). When visualized in a gene-expression heatmap across the GTEx v8 tissues, some of the genes, including *EBF1, GNAQ, HAND2, ZBTB38*, and *COL27A1*, clustered in a profile of higher expression in tissues including blood vessel, uterus, and fallopian tube (Figure S5). Gene set analysis based on a list of genes corresponding to all significant loci in the current study identified enrichment of multiple pathways, with GO terms referring to regulation of morphogenesis and development of various organs and tissues among multiple top pathways (Table S6).

### Replication and joint analysis

We used data from the Nordic data sets to test for replication of the associated loci and to perform joint analysis (Table S7). Loci near *WNT4, EEFSEC, EBF1*, and *AGTR2* were not included, since the same replication data was used in the study that discovered these associations^8^. While all effects among the genome-wide significant meta-analysis loci and the replication population were in the same direction, the strongest associations in the replication population were detected for *ZBTB38, HAND2, TET3*, and *KCNAB1*. Joint analysis of the replication data and the meta-analysis variants with suggestive significance (*p*<1e−6 to 5e−8) detected *DNAH2* and *RAP2C* as additional loci associated with gestational duration. In addition, the current meta-analysis detected many of the associations in the preprint reporting maternal meta-analysis of the timing of parturiton^10^, including nine loci for gestational duration and two for SPTB.

### Characterization of association signals

To gain insight into the nature of the associated loci, we explored previous associations with other complex traits in the literature and with data from FinnGenR7 and the IEU openGWAS project^13^, and performed colocalization analysis with expression quantitative trait loci (eQTLs) to evaluate if the associated variants affect their target genes by regulating gene expression. In the FinnGen data, we screened the meta-analysis lead variants for associations with all >3000 phenotypes in freeze 7 (Figure S8), whereas data from the IEU openGWAS project was queried in a PheWAS for all associated variants within the associated meta-analysis loci (Table S8). The lead variant rs1991431 in *ZBTB38* with a replicable association was associated with hyperplasia of prostate (BHP) in the FinnGen (Figure S8), and other associated variants in the locus were linked to various complex traits including cell counts of lymphocytes and monocytes, and *ZBTB38* mRNA expression on the IEU openGWAS data. The same alleles of several meta-analysis variants (e.g., T allele of variant rs9846396 associated with longer gestational duration; *Z*-score = 6.36, *p* = 2.04e−10) were also associated with taller height, higher body mass measures, and increased risk of prostate cancer (Table S8). We detected colocalization for variants associated with gestational duration and *ZBTB38* expression in tissues including monocytes, T cells, and B cells, and alleles associated with longer gestational duration were linked to higher *ZBTB38* expression (Figure 3, Table S9).

**Figure 3.**
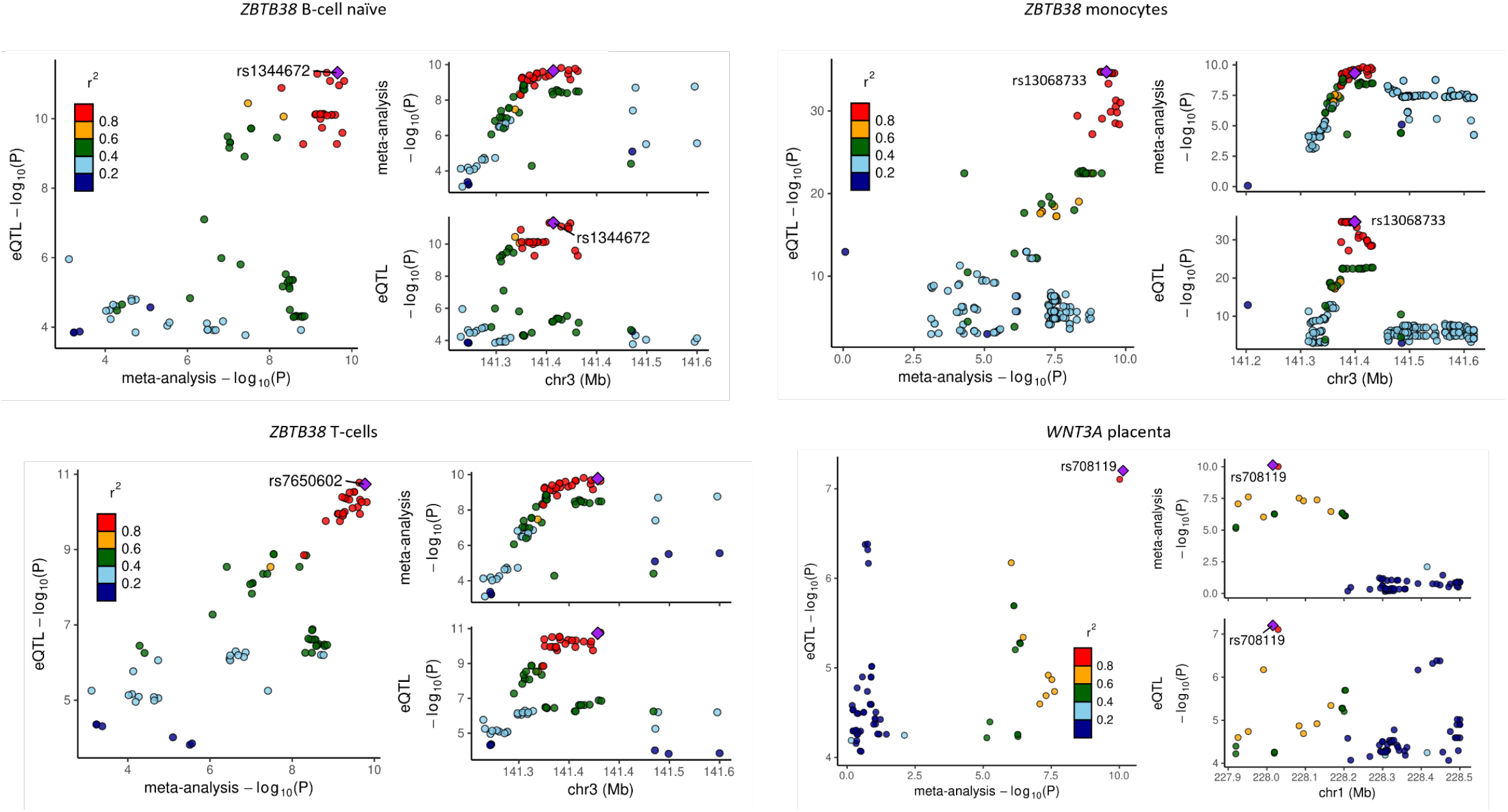
Colocalization analysis of meta-analysis associations with expression quantitative trait (eQTL) data. The variants are colored according to their LD (r^2^) with the lead SNP, based on pairwise LD in European population of the 1000 Genomes Project Phase 3. **A–C)** *ZBTB38* variants were implicated in gestational duration-linked gene regulation in various immune regulatory cell types. **D)** Variants in *WNT3A* locus colocalized with *WNT3A* expression in placenta.

Like variants in *ZBTB38*, the polymorphisms in the *WNT3A* locus were associated with height and body mass indices, and the encoded protein was implicated in cell fate and patterning during embryogenesis^14,15^. Concordantly, variants associated with gestational duration colocalized with *WNT3A* expression in the placenta (Figure3, Table S9).

We detected a replicable association of variants near *HAND2. HAND2* plays a role in heart development with previous associations with traits including atrial fibrillation and platelet count^11^. *HAND2* is expressed in the human uterine tissue, where it is upregulated by the progesterone receptor, and involved in immune tolerance of the decidua by regulating a distinct set of genes including interleukin 15^16,17^.

*TET3* and *KCNAB1* showed strong replication (Table S7). Variants in both genes were associated with birth weight of offspring (Table S8), and alleles associated with longer gestational duration in the current meta-analysis were linked to higher birth-weight measures. We observed a similar positive correlation between gestational duration and birth weight at the genome-wide level in the LDSC analysis (Figure 2). Variants associated with gestational duration colocalized with *KCNA1B* expression two reproductive tissues and blood vessel (Table S9). Alleles linked to longer gestational duration were associated with increased *KCNAB1* expression in all tissues.

The case-control meta-analysis of SPTB detected two novel associated loci: *GC* and *LINC02824*, which encode vitamin D-binding protein and a long noncoding RNA, respectively. *GC* is involved in vitamin D transport and storage, and circulating vitamin D levels have been linked to preterm birth and other pregnancy- and reproductive health–related outcomes in observational studies^18,19^.

## Discussion

The current genome-wide meta-analysis of SPTB (n=98,371) and gestational duration (n=68,732) identified several associated loci. We detected loci that had no previous associations with gestational duration or SPTB, and our findings further reinforce the associations of genes from previous GWASs of mothers who gave birth preterm. The associated loci with strong replication in the current analysis, including *ZBTB38, HAND2, TET3*, and *KCNAB1*, also showed association in the recent meta-analysis of the timing of parturition^10^, establishing these genes as strong candidates for further molecular biological studies of SPTB. The inferred functions of the assigned candidate genes were consistent with a role in the timing of birth.

Association of *ZBTB38*, zinc finger and BTB domain containing 38, with benign hyperplasia of prostate (BHP) is a compelling finding given that both gestational duration and BHP are affected by changes in estrogen and androgen levels^20,21^. *ZBTB38* was further associated with cell counts of various immune cells, and our results suggest that increased *ZBTB38* expression in these cell types may play a role in regulating length of pregnancy. Alleles associated with longer gestational duration showed association with increased height, body mass, and risk of prostate cancer. It remains to be determined whether *ZBTB38* confers its effect on birth timing through pregnancy-specific mechanisms or by contributing to more general immune pathways that influence gestation. Our findings for *ZBTB38* associations are in keeping with reported correlations among maternal height, gestational duration, and fetal growth, and further comply with detected associations between maternal birth weight–elevating alleles and longer gestational duration and between maternal gestation-prolonging alleles and the risk of prostate carcinoma^8,22,23^.

An association near *HAND2* showed strong replication. *HAND2* encodes heart and neural crest derivatives expressed 2, a transcription factor best known for its roles in cardiac morphogenesis and limb development. Decreasing expression of *HAND2* in the decidua during pregnancy may contribute to regulation of gestational duration^24^. The expression of *HAND2* in the human uterine tissue, and its gradually decreasing expression in the decidua during pregnancy^24^, makes it an interesting candidate gene and a potential biomarker for SPTB.

Variants in Tet methylcytosine dioxygenase 3 *(TET3)* and potassium voltage-gated channel subfamily A regulatory beta subunit 1 (*KCNAB1)* were previously associated with birth weight of offspring, and alleles associated with longer gestational duration correlated with birth-weight measures, complying with known correlations of the length of gestation and fetal growth^22^. It is possible that the associations between these loci and gestational duration explain the effect of the mentioned loci on birth weight. Our results further suggest that *KCNAB1* expression contributes to the timing of birth. Of note, *TET3* was suggested to play a role in embryo implantation^25^. Both *TET3* and *KCNAB1* represent interesting targets for further study to determine their specific roles related to the regulation of gestational duration.

Interestingly, *COL27A1* was associated with phenotypes including embryonic growth retardation, abnormal placenta morphology, and abnormal placenta vasculature in data in the knock-out mice as investigated via IMPC (https://www.mousephenotype.org/). *COL27A1* is most abundantly expressed in the endometrium, and the gene encodes collagen type XXVII alpha 1 chain, which is a fibrillar, developmentally regulated protein. Further, the meta-analysis lead variant of the *COL27A1* locus is near miR-455, which has roles in cartilage development, adipogenesis, and preeclampsia, and may protect endometrial cells against oxidative stress^26,27^.

The novel meta-analysis loci associated with gestational duration, such as variants in *GNAQ* and *KCNN2*, comprised further intriguing candidates. *GNAQ* (protein subunit alpha q) plays a role in survival of immune cells and was identified as part of a transcriptomic signature related to human labor in the choriodecidua^28,29^. *KCNN2* (potassium calcium-activated channel subfamily N member 2) has no obvious connection to pregnancy-related regulation, but *KCNN3*, another molecule in the KCNN family of potassium channel genes, plays a role in uterine function^30^. The role of these risk loci and corresponding causal genes remains to be verified.

The case-control meta-analysis of SPTB detected an association in *GC*, encoding GC vitamin D binding protein, as of special interest because of its known involvement in vitamin D transport and storage. Protein encoded by *GC* is the primary carrier of vitamin D that binds to the vitamin and its plasma metabolites and transports them to their target tissues. Previous studies have suggested links among plasma vitamin D levels and preterm birth or other pregnancy-related outcomes including pre-eclampsia, polycystic ovary syndrome, and endometriosis^18,19^. Vitamin D deficiency was found associated with many adverse outcomes including those related to pregnancy, whereas increased levels of the protein product of *GC* showed association with a reduced risk of certain immune-mediated diseases^19,31,32^. The precise role of *GC* in the context of human pregnancy and SPTB remains to be determined.

Altogether, our results highlight the unique nature of SPTB. At the genome-wide level, birth weight measures were the only traits that showed significant correlations with both gestational duration and SPTB in a comprehensive set of complex phenotypes. However, the Bonferroni correction deployed for the 773 tests is likely overly conservative since the traits include closely related phenotypes. Gene set enrichment for gestational duration and pathway analysis for gestational duration and SPTB highlighted involvement of gonad development and steroid hormone biosynthetic processes and GO terms referring to regulation of morphogenesis among multiple top pathways. Gene set enrichment analysis of SPTB implied kinetochore microtubule as the top pathway. Proper function of the kinetochore-microtubule pathway is essential for preserving genomic integrity and prevention of birth defects^33^. Tissue analysis pinpointed several reproductive tissues, including uterus and ovary, among the most relevant tissue types for both SPTB- and gestational duration-associated genes.

Multiple variants in the candidate loci were individually associated with birth weight indices. Associated genes had primary roles in steroid hormone–regulating processes and tissue and organ morphogenesis. Reproductive tissues of the mother were among the principal locations where the associated genes were expressed. Our results suggest that many of the associated variants contribute to pregnancy outcomes by regulating expression of their target genes, mainly but not exclusively in reproductive tissues and immune cell types. Hence, our results indicate that those tissue and cell types are the most relevant when considering the regulatory events related to pregnancy and preterm birth, and should be the primary targets in future molecular biological studies of SPTB and gestational duration.

The current analysis was restricted to individuals of predominantly European descent. Future studies should include ancestrally diverse populations to better understand the genetic architecture of the timing of birth and to ensure the broad applicability of results from genetic studies^34^.

In conclusion, the current meta-analysis detected multiple loci that were associated with gestational duration or SPTB and produced intriguing candidates for further studies. Our results highlight the intricate nature of spontaneous birth as a trait and emphasize the importance of reproductive and immune tissues and cell types. The new genetic discoveries prime further research including large-scale complex investigations and individual regulatory pathway analyses utilizing labor-inducing tissues and cells. Studies may eventually reveal signaling pathways that activate spontaneous preterm birth and contribute towards effective prevention of SPTB.

## Methods

### Study cohorts, phenotype descriptions, and ethics information

We conducted a meta-analysis of SPTB (8,542 cases and 89,829 controls) and a quantitative meta-analysis of gestational duration (n=68,733) with data from mothers of European ancestry. The data originated from FinnGen, Northern/Central Finland, and 23andMe project.

**The FinnGen research project** (launched 2017) combines genome information with health care data from national registries. The project aims to collect data from 500,000 Finnish participants. Preterm and term birth were defined as births before and after 37 weeks of gestation. FinnGen preterm endpoint in Preparatory Phase Data Freeze 6 comprised individuals with World Health Organization International Classification of Diseases, Eight, Ninth, and Tenth Revision (ICD-8, ICD-9, and ICD-10) codes O60, 644, and 63497, respectively. We excluded births with ICD-9 code 644 (“early or threatened labor”) if they occurred after 37 weeks of gestation according to birth register data, and individuals with multiple gestation or birth, preeclampsia/eclampsia, and polyhydroamnios. Controls were people with spontaneous term birth. The GWAS of SPTB comprised 4,925 cases and 49,105 controls, and the GWAS of gestational duration comprised 24,391 mothers for whom gestational duration was available. We additionally performed GWAS with a “strict” definition of SPTB, in which we only included cases with births indicated as spontaneous and preterm in the endpoint data.

FinnGen participants provided informed consent under the Finnish Biobank Act. Older cohorts with study-specific consents were transferred to the Finnish biobanks after approval by Fimea, the National Supervisory Authority for Welfare and Health. Recruitment protocols followed the biobank protocols approved by Fimea. The Coordinating Ethics Committee of the Hospital District of Helsinki and Uusimaa (HUS) approved the FinnGen study protocol (Nr HUS/990/2017).

The FinnGen study is approved by the Finnish Institute for Health and Welfare (permit numbers THL/2031/6.02.00/2017, THL/1101/5.05.00/2017, THL/341/6.02.00/2018, THL/2222/6.02.00/2018, THL/283/6.02.00/2019, THL/1721/5.05.00/2019, THL/1524/5.05.00/2020, and THL/2364/14.02/2020), the Digital and Population Data Service Agency (permit numbers VRK43431/2017-3, VRK/6909/2018-3, and VRK/4415/2019-3), the Social Insurance Institution (permit numbers KELA 58/522/2017, KELA 131/522/2018, KELA 70/522/2019, KELA 98/522/2019, KELA 138/522/2019, KELA 2/522/2020, and KELA 16/522/2020), and Statistics Finland (permit numbers TK-53-1041-17 and TK-53-90-20). The Biobank Access Decisions for FinnGen samples and data utilized in FinnGen Data Freeze 6 include: THL Biobank BB2017_55, BB2017_111, BB2018_19, BB_2018_34, BB_2018_67, BB2018_71, BB2019_7, BB2019_8, BB2019_26, BB2020_1, Finnish Red Cross Blood Service Biobank 7.12.2017, Helsinki Biobank HUS/359/2017, Auria Biobank AB17-5154, Biobank Borealis of Northern Finland_2017_1013, Biobank of Eastern Finland 1186/2018, Finnish Clinical Biobank Tampere MH0004, Central Finland Biobank 1-2017, and Terveystalo Biobank STB 2018001.

The study subjects from **Northern and Central Finland** were sampled in Oulu and Tampere University Hospital districts. Informed consent was obtained from the mother. SPTB was defined as birth prior to 36 wk + 1 d of gestation. Term birth was defined as birth at 38–41 wk (38 wk + 0 d to 41 wk + 6 d) of gestation. We excluded births with multiple gestation, preeclampsia, polyhydroamnios, intrauterine growth restriction, placental abruption, anomalies of the fetus, clinical chorioamnionitis or acute septic infection in the mother, alcohol or narcotic use, and accidents. Term births were from families without previous preterm births. The analysis comprised 286 cases and 488 controls. Ethical approval was received from the participating centers (Oulu University Hospital 79/2003, 14/2010, and 73/2013).

Summary statistics of the **23andMe research program** were obtained by request. Women included in the analyses provided informed consent and answered surveys online in accordance with a human subjects protocol approved by Ethical & Independent Review Services (http://www.eandireview.com). The summary data comprised unrelated mothers of European ancestry with self-reported gestational duration for their first singleton live birth. Individuals reporting a medical indication for preterm delivery were excluded. Preterm birth was defined as birth before 37 weeks of gestation, and control samples were people with term delivery. The meta-analysis included 43,568 individuals (3331 cases and 40,237 controls).

Data used in the replication and joint analysis originated from European women in the **Nordic data sets** including FIN cohort (N=888; Finland), MoBa (N=1,834; Norway), and DNBC (N=5,921; Denmark), for which the summary statistics were obtained via collaboration^8,35,36^. To protect individual level data, we show distribution of gestational duration in the Finnish studies as categories (Table S1). Other cohorts were described previously^8,35,36^.

### DNA sample preparation, genotyping, imputation, and quality control

Various methods were used to extract DNA from the FinnGen samples. Genotyping was done with Illumina and Affymetrix arrays (Illumina Inc, San Diego, CA and Thermo Fisher Scientific, Santa Clara, CA). Sample quality control (QC) entailed excluding individuals of uncertain sex, non-Finnish ancestry, high missingness (>5%), and excess heterozygosity (±4SD). For genotype QC, variants with missingness >2%, minor allele count (MAC)<3, and deviation from Hardy–Weinberg equilibrium (HWE) (*p*<1e−6) were excluded. Imputation was conducted against a Finnish population–specific SISuv3 reference with Beagle4.1^37^. Variants with imputation info (INFO)<0.7 were excluded (https://github.com/FINNGEN/finngen-documentation).

DNA from Norhern/Central Finnish study was extracted with UltraClean Blood DNA Isolation Kit (MO BIO Laboratories, Inc., Carlsbad, CA), Puregene Blood Core Kit (Qiagen, Hilden, Germany), or prepIT-L2P kit (DNA Genotek, Ontario, Canada). Genotyping was performed with the Infinium HumanCoreExome BeadChip (Illumina, San Diego, CA) by the Technology Centre, Institute for Molecular Medicine Finland (FIMM), University of Helsinki. Variants with minor allele frequency (MAF)<1%, HWE *p*<1e-4, or genotyping rate <90%, and samples with >10% missingness, were excluded. Prephasing was conducted with SHAPEIT2^38^, andimputation with IMPUTE2^39^, against the 1000 Genomes Project (1KGP) v3 reference panel^40^. Variants with INFO<0.7 were excluded.

DNA of the 23andMe samples was extracted from saliva samples, followed by genotyping with custom Illumina platforms by the National Genetics Institute (NGI). Samples with <97% European ancestry, and variants with HWE *p*<10e-20, call rate <95%, or allele frequency discrepancy with 1KGP Europeans, were excluded. Imputation was done with Minimac2^42^, using the 1KGP phase1^40^.

### GWAS and meta-analysis

FinnGen GWAS was conducted with Scalable and Accurate Implementation of GEneralized mixed model (SAIGE)^41^. Gestational duration were inverse normalized. GWAS covariates were age, sex, genotyping batch, and ten leading principal components. MAC was set to five.

We used SNPTESTv2^42^ in GWAS of Northern/Central Finnish cohort. A frequentist case– control association test was implemented for SPTB, and a quantitative trait test for gestational duration was carried out with a linear model. Covariates were three multidimensional scaling dimensions, defined with Plink1.9^43^. We used SNPTEST defaults to achieve mean centering and scaling of gestational duration and to apply quantile normalization. Post-GWAS QC entailed excluding variants with MAF<1% and SNPTEST info<0.7.

In the 23andMe data, preterm birth was analyzed with logistic regression, and linear regression was applied in the GWAS of gestational duration. Covariates were maternal age and the top five principal components.

We used METAL^44^ to conduct a fixed-effect inverse variance–weighted meta-analysis of SPTB and a sample size–weighted *p*-value–based meta-analysis of gestational duration. Sample size-based meta-analysis allows combining results when β-coefficients and standard errors from individual studies are in different units. Genomic coordinates of the meta-analysis cohorts were aligned into the GRCh38 coordinates. Genomic inflation factor was calculated with linkage disequilibrium score regression (LDSC)^12^.We report associations based on at least two individual meta-analysis cohorts, and excluded remaining rare variants with MAF<0.01%.

### Characterization of association signals

We defined associated loci as genomic regions within a ±1 Mb window around the lead variant. The locus was defined as novel if there were no previous genome-wide significant associations for SPTB or gestational duration in the ±1 Mb window in the National Human Genome Research Institute–European Bioinformatics Institute (NHGRI-EBI) GWAS Catalog or in the preprint of meta-analysis of the timing of parturition^10,11^.

We used LDSC to estimate SNP-based heritability and to test for genetic correlation between gestational duration or SPTB with a comprehensive set of phenotypes downloaded from the Integrative Epidemiology Unit (IEU) OpenGWAS Project^12,13^. We used FUMA GWAS (Functional Mapping and Annotation of Genome-Wide Association Studies)^45^ to aid functional annotation of the GWAS results. FUMA was used to prioritize genes for enrichment testing and assessment and visualization of tissue-specific expression among GTExv8 tissues^46^. FUMA implements MAGMA (Multi-marker Analysis of GenoMic Annotation)^47^ in gene-based analyses and gene-set enrichment analyses for GWAS summary data with curated gene sets and GO terms from Molecular Signature Database, MSigDB. In addition, we tested lists of gestational duration- and STPB-associated candidate genes for averaged gene expression across GTExv8 tissues (hierarchical clustering) and for enrichment against various gene sets with hypergeometric tests in FUMA’s GENE2FUNC process.

To gain insight into the associated loci, we checked previous associations in the FinnGen R7 data and performed phenome-wide association study (PheWAS) within 1 Mb window around the meta-analysis index variants by querying GWAS data in the IEU OpenGWAS Project, which includes approximately 40,000 studies^13^.

We performed colocalization analysis with HyprColoc^48^ to assess if associated variants were also quantitative trait loci (QTLs) that affect mRNA expression. Colocalization was tested for variants within a 1Mb window around the meta-analysis lead variant. We estimated betas from the meta-analysis *Z*-scores^49^. We used expression QTLs (eQTLs) from the eQTLCatalogue^50^, which contains uniformly processed cis-eQTLs from most of the available public studies. From the GTEx data in the eQTLCatalogue, we included eQTLs based on GTEx v8 and LCLs from an earlier release. We report colocalization results with a posterior probability (PP)>0.6 and meta-analysis *p*<5e-8. We did not test eQTLs with FDR>0.05.

### Replication and joint analysis

We tested the lead variants within each locus for association in the replication data from the data set of Nordic birth studies. For most of the tested variants, replication data from two out of three cohorts (FIN and MoBa, N=2,722) was available. Joint test of the replication data and meta-analysis variants with *p*<1e-6 was done to see if additional loci reached genome-wide significance. We did not test replicated loci originally reported in Zhang et al.^8^since the same replication data was used.

## Data Availability

Meta-analysis summary statistics of the top 10,000 SNPs will be deposited in an appropriate data repository. Summary statistics of the Northern/Central Finnish cohort are available upon reasonable request. The full GWAS summary statistics for the 23andMe discovery data set will be made available through 23andMe to qualified researchers under an agreement with 23andMe that protects the privacy of the 23andMe participants. Please visit https://research.23andme.com/collaborate/#dataset-access/ for more information and to apply to access the data. For information regarding FinnGen data access, please visit www.finngen.fi.

https://www.finngen.fi/

## Funding

The FinnGen project is funded by two grants from Business Finland (HUS 4685/31/2016 and UH 4386/31/2016) and the following industry partners: AbbVie Inc., AstraZeneca UK Ltd, Biogen MA Inc., Bristol Myers Squibb (and Celgene Corporation & Celgene International II Sàrl), Genentech Inc., Merck Sharp & Dohme Corp, Pfizer Inc., GlaxoSmithKline Intellectual Property Development Ltd., Sanofi US Services Inc., Maze Therapeutics Inc., Janssen Biotech Inc, Novartis AG, and Boehringer Ingelheim. Research regarding spontaneous preterm birth performed at the University of Oulu was financed by the Jane and Aatos Erkko Foundation (MH, MR), Competitive State Research Financing of the Expert Responsibility Area of Oulu University Hospital (MR), Sigrid Jusélius Foundation (MH), Foundation for Pediatric Research (MR), Emil Aaltonen Foundation (AP), and Alma and K.A. Snellman Foundation (AP). BF received support from an Oak Foundation fellowship and a Novo Nordisk Foundation grant (12955). GZ is supported by the Eunice Kennedy Shriver National Institute of Child Health & Human Development of the National Institutes of Health under Award Number R01HD101669, the Burroughs Wellcome Fund (Grant 10172896), the March of Dimes Prematurity Research Center Ohio Collaborative and the Bill & Melinda Gates Foundation. The Norwegian Mother and Child Cohort Study was supported by the Norwegian Ministry of Health and the Ministry of Education and Research, by the National Institute of Environmental Health Sciences (contract no. N01-ES-75558), the National Institute of Neurological Disorders and Stroke (UO1 NS 047537-01 and UO1 NS 047537-06A1), the Norwegian Research Council/FUGE (151918/S10, 183220/S10 and FRI-MEDBIO 249779). BJ was funded by the Swedish Research Council (2015-02559) and by from the Norwegian Research Council, a grant from the Jane and Dan Olsson Foundations, a grant (ALFGBG-426411) from the Swedish government to researchers in the public health service. The Danish National Birth Cohort was established with a significant grant from the Danish National Research Foundation. Additional support was obtained from the Danish Regional Committees, the Pharmacy Foundation, the Egmont Foundation, the March of Dimes Birth Defects Foundation, the Health Foundation and other minor grants. The DNBC Biobank has been supported by the Novo Nordisk Foundation and the Lundbeck Foundation.

## Acknowledgements

We want to acknowledge the participants and investigators of the FinnGen study. The following biobanks are acknowledged for delivering biobank samples to FinnGen: Auria Biobank (www.auria.fi/biopankki), THL Biobank (www.thl.fi/biobank), Helsinki Biobank (www.helsinginbiopankki.fi), Biobank Borealis of Northern Finland (https://www.ppshp.fi/Tutkimus-ja-opetus/Biopankki/Pages/Biobank-Borealis-briefly-in-English.aspx), Finnish Clinical Biobank Tampere (www.tays.fi/en-US/Research_and_development/Finnish_Clinical_Biobank_Tampere), Biobank of Eastern Finland (www.ita-suomenbiopankki.fi/en), Central Finland Biobank (www.ksshp.fi/fi-FI/Potilaalle/Biopankki), Finnish Red Cross Blood Service Biobank (www.veripalvelu.fi/verenluovutus/biopankkitoiminta), and Terveystalo Biobank (www.terveystalo.com/fi/Yritystietoa/Terveystalo-Biopankki/Biopankki/). All Finnish Biobanks are members of BBMRI.fi infrastructure (www.bbmri.fi). Finnish Biobank Cooperative–FINBB (https://finbb.fi/) is the coordinator of BBMRI-ERIC operations in Finland. Finnish biobank data can be accessed through Fingenious® services (https://site.fingenious.fi/en/), managed by FINBB. We would like to thank the research participants and employees of 23andMe, Inc, for making this work possible. CSC–IT Center for Science, Finland, is acknowledged for computational resources. This study includes data from the Norwegian Mother, Father and Child Cohort Study (MoBa) conducted by the Norwegian Institute of Public Health and from the Danish National Birth Cohort (DNBC), and we would like to thank the research participants of the Norwegian MoBa study and the DNBC. We thank Maarit Haarala (University of Oulu, Oulu, Finland) and Riitta Vikeväinen (Oulu University Hospital, Oulu, Finland) for technical assistance.

## Data availability statement

Meta-analysis summary statistics of the top 10,000 SNPs will be deposited in an appropriate data repository. Summary statistics of the Northern/Central Finnish cohort are available upon reasonable request. The full GWAS summary statistics for the 23andMe discovery data set will be made available through 23andMe to qualified researchers under an agreement with 23andMe that protects the privacy of the 23andMe participants. Please visit https://research.23andme.com/collaborate/#dataset-access/ for more information and to apply to access the data. For information regarding FinnGen data access, please visit https://www.finngen.fi.

## Notes

### Competing Interest Statement

The authors have declared no competing interest.

### Author Declarations

Ethical permissions of the used FinnGen data release: FinnGen participants provided informed consent under the Finnish Biobank Act. Older cohorts with study-specific consents were transferred to the Finnish biobanks after approval by Fimea, the National Supervisory Authority for Welfare and Health. Recruitment protocols followed the biobank protocols approved by Fimea. The Coordinating Ethics Committee of the Hospital District of Helsinki and Uusimaa (HUS) approved the FinnGen study protocol (Nr HUS/990/2017). The FinnGen study is approved by the Finnish Institute for Health and Welfare (permit numbers THL/2031/6.02.00/2017, THL/1101/5.05.00/2017, THL/341/6.02.00/2018, THL/2222/6.02.00/2018, THL/283/6.02.00/2019, THL/1721/5.05.00/2019, THL/1524/5.05.00/2020, and THL/2364/14.02/2020), the Digital and Population Data Service Agency (permit numbers VRK43431/2017-3, VRK/6909/2018-3, and VRK/4415/2019-3), the Social Insurance Institution (permit numbers KELA 58/522/2017, KELA 131/522/2018, KELA 70/522/2019, KELA 98/522/2019, KELA 138/522/2019, KELA 2/522/2020, and KELA 16/522/2020), and Statistics Finland (permit numbers TK-53-1041-17 and TK-53-90-20). The Biobank Access Decisions for FinnGen samples and data utilized in FinnGen Data Freeze 6 include: THL Biobank BB2017_55, BB2017_111, BB2018_19, BB_2018_34, BB_2018_67, BB2018_71, BB2019_7, BB2019_8, BB2019_26, BB2020_1, Finnish Red Cross Blood Service Biobank 7.12.2017, Helsinki Biobank HUS/359/2017, Auria Biobank AB17-5154, Biobank Borealis of Northern Finland_2017_1013, Biobank of Eastern Finland 1186/2018, Finnish Clinical Biobank Tampere MH0004, Central Finland Biobank 1-2017, and Terveystalo Biobank STB 2018001. Nothern/Central Finnish cohort: Ethics committee of Pohjois-Pohjanmaan sairaanhoitopiirin kuntayhtyma gave ethical approval for this work. Summary statistics of the 23andMe research program were obtained by request. Women included in the analyses provided informed consent and answered surveys online in accordance with a human subjects protocol approved by Ethical & Independent Review Services (http://www.eandireview.com).

## References

1. Goldenberg, R. L., Culhane, J. F., Iams, J. D. & Romero, R. Epidemiology and causes of preterm birth. Lancet 371, 75–84 (2008).

2. Blencowe, H. et al. Born Too Soon: The global epidemiology of 15 million preterm births. Reprod Health 10, 1–14 (2013).

3. Jakobsson, M., Gissler, M., Paavonen, J. & Tapper, A. M. The incidence of preterm deliveries decreases in Finland. BJOG 115, 38–43 (2008).

4. Wikström, T. et al. Effect of second-trimester sonographic cervical length on the risk of spontaneous preterm delivery in different risk groups: A prospective observational multicenter study. Acta Obstet Gynecol Scand 100, 1644–1655 (2021).

5. Boyd, H. A. et al. Maternal Contributions to Preterm Delivery. Am J Epidemiol 170, 1358–1364 (2009).

6. York, T. P. et al. Fetal and maternal genes’ influence on gestational age in a quantitative genetic analysis of 244,000 Swedish births. Am J Epidemiol 178, 543–550 (2013).

7. Plunkett, J. et al. Mother’s Genome or Maternally-Inherited Genes Acting in the Fetus Influence Gestational Age in Familial Preterm Birth. Hum Hered 68, 209 (2009).

8. Zhang, G. et al. Genetic Associations with Gestational Duration and Spontaneous Preterm Birth. New England Journal of Medicine 377, 1156–1167 (2017).

9. Liu, X. et al. Variants in the fetal genome near pro-inflammatory cytokine genes on 2q13 associate with gestational duration. Nature Communications 2019 10:1 10, 1–13 (2019).

10. Solé-Navais, P. et al. Genetic effects on the timing of parturition and links to fetal birth weight. medRxiv 32, 2022.05.04.22274624 (2022).

11. J, M. et al. The new NHGRI-EBI Catalog of published genome-wide association studies (GWAS Catalog). Nucleic Acids Res 45, D896–D901 (2017).

12. Bulik-Sullivan, B. et al. LD Score regression distinguishes confounding from polygenicity in genome-wide association studies. Nat Genet 47, 291–295 (2015).

13. Elsworth, B. et al. The MRC IEU OpenGWAS data infrastructure. bioRxiv 2020.08.10.244293 (2020) doi:10.1101/2020.08.10.244293.

14. Minn, K. T., Dietmann, S., Waye, S. E., Morris, S. A. & Solnica-Krezel, L. Gene expression dynamics underlying cell fate emergence in 2D micropatterned human embryonic stem cell gastruloids. Stem Cell Reports 16, 1210 (2021).

15. Takada, S. et al. Wnt-3a regulates somite and tailbud formation in the mouse embryo. Genes Dev 8, 174–189 (1994).

16. Murata, H., Tanaka, S. & Okada, H. Immune Tolerance of the Human Decidua. J Clin Med 10, 1–16 (2021).

17. Sakabe, N. J. et al. Transcriptome and regulatory maps of decidua-derived stromal cells inform gene discovery in preterm birth. Sci Adv 6, (2020).

18. Kiely, M. E., Wagner, C. L. & Roth, D. E. Vitamin D in pregnancy: Where we are and where we should go. J Steroid Biochem Mol Biol 201, (2020).

19. Fernando, M. et al. Vitamin D-Binding Protein in Pregnancy and Reproductive Health. Nutrients 12, (2020).

20. Makieva, S., Saunders, P. T. K. & Norman, J. E. Androgens in pregnancy: roles in parturition. Hum Reprod Update 20, 542 (2014).

21. Nicholson, T. M. & Ricke, W. A. Androgens and estrogens in benign prostatic hyperplasia: Past, present and future. Differentiation 82, 184–199 (2011).

22. Beaumont, R. N. et al. Genome-wide association study of offspring birth weight in 86 577 women identifies five novel loci and highlights maternal genetic effects that are independent of fetal genetics. Hum Mol Genet 27, 742 (2018).

23. Gudmundsson, J. et al. Genome-wide association and replication studies identify four variants associated with prostate cancer susceptibility. Nat Genet 41, 1122 (2009).

24. Marinić, M., Mika, K., Chigurupati, S. & Lynch, V. J. Evolutionary transcriptomics implicates hand2 in the origins of implantation and regulation of gestation length. Elife 10, 1–52 (2021).

25. Liu, A. et al. Loss of miR-29a impairs decidualization of endometrial stromal cells by TET3 mediated demethylation of Col1A1 promoter. iScience 24, (2021).

26. Tang, W., Chen, O., Yao, F. & Cui, L. miR-455 targets FABP4 to protect human endometrial stromal cells from cytotoxicity induced by hydrogen peroxide. Mol Med Rep 20, 4781–4790 (2019).

27. Swingler, T. E. et al. The expression and function of microRNAs in chondrogenesis and osteoarthritis. Arthritis Rheum 64, 1909–1919 (2012).

28. Bhattacharya, S. et al. Lymphocyte-Specific Biomarkers Associated With Preterm Birth and Bronchopulmonary Dysplasia. Front Immunol 11, 1 (2021).

29. Lui, S. et al. Delineating differential regulatory signatures of the human transcriptome in the choriodecidua and myometrium at term labor,. Biol Reprod 98, 422–436 (2018).

30. Lu, Y. C. et al. Small-conductance, calcium-activated potassium channel 3 (SK3) is a modulator of endometrial remodeling during endometrial growth. J Clin Endocrinol Metab 99, 3800– 3810 (2014).

31. Mansur, J. L., Oliveri, B., Giacoia, E., Fusaro, D. & Costanzo, P. R. Vitamin D: Before, during and after Pregnancy: Effect on Neonates and Children. Nutrients 14, (2022).

32. Albiñana, C. et al. Genetic correlates of vitamin D-binding protein and 25 hydroxyvitamin D in neonatal dried blood spots. doi:10.1101/2022.06.08.22276164.

33. Weaver, B. A. A. & Cleveland, D. W. Decoding the links between mitosis, cancer, and chemotherapy: The mitotic checkpoint, adaptation, and cell death. Cancer Cell 8, 7–12 (2005).

34. Peterson, R. E. et al. Genome-wide Association Studies in Ancestrally Diverse Populations: Opportunities, Methods, Pitfalls, and Recommendations. Cell 179, 589–603 (2019).

35. Zhang, G. et al. Assessing the Causal Relationship of Maternal Height on Birth Size and Gestational Age at Birth: A Mendelian Randomization Analysis. PLoS Med 12, (2015).

36. Magnus, P. et al. Cohort Profile Update: The Norwegian Mother and Child Cohort Study (MoBa). Int J Epidemiol 45, 382–388 (2016).

37. Browning, B. L. & Browning, S. R. Genotype Imputation with Millions of Reference Samples. Am J Hum Genet 98, 116–126 (2016).

38. Delaneau, O., Marchini, J. & Zagury, J. F. A linear complexity phasing method for thousands of genomes. Nat Methods 9, 179–181 (2011).

39. Howie, B. N., Donnelly, P. & Marchini, J. A flexible and accurate genotype imputation method for the next generation of genome-wide association studies. PLoS Genet 5, (2009).

40. Auton, A. et al. A global reference for human genetic variation. Nature 2015 526:7571 526, 68–74 (2015).

41. Zhou, W. et al. Efficiently controlling for case-control imbalance and sample relatedness in large-scale genetic association studies. Nat Genet 50, 1335 (2018).

42. Marchini, J., Howie, B., Myers, S., McVean, G. & Donnelly, P. A new multipoint method for genome-wide association studies by imputation of genotypes. Nature Genetics 2007 39:7 39, 906–913 (2007).

43. Chang, C. C. et al. Second-generation PLINK: Rising to the challenge of larger and richer datasets. Gigascience 4, 7 (2015).

44. Willer, C. J., Li, Y. & Abecasis, G. R. METAL: fast and efficient meta-analysis of genomewide association scans. Bioinformatics 26, 2190 (2010).

45. Watanabe, K., Taskesen, E., van Bochoven, A. & Posthuma, D. Functional mapping and annotation of genetic associations with FUMA. Nature Communications 2017 8:1 8, 1–11 (2017).

46. Aguet, F. et al. The GTEx Consortium atlas of genetic regulatory effects across human tissues. bioRxiv 787903 (2019) doi:10.1101/787903.

47. de Leeuw, C. A., Mooij, J. M., Heskes, T. & Posthuma, D. MAGMA: Generalized Gene-Set Analysis of GWAS Data. PLoS Comput Biol 11, e1004219 (2015).

48. Foley, C. N. et al. A fast and efficient colocalization algorithm for identifying shared genetic risk factors across multiple traits. Nature Communications 2021 12:1 12, 1–18 (2021).

49. Zhu, Z. et al. Integration of summary data from GWAS and eQTL studies predicts complex trait gene targets. Nature Genetics 2016 48:5 48, 481–487 (2016).

50. Kerimov, N. et al. A compendium of uniformly processed human gene expression and splicing quantitative trait loci. Nature Genetics 2021 53:9 53, 1290–1299 (2021).

