## Supplementary figures 1-8 for "Meta-analysis of gestational duration and spontaneous preterm birth identifies new maternal risk loci"

[3]Contributors of FinnGen are listed in supplemental material

[4]Division of Human Genetics, Cincinnati Children's Hospital Medical Center, Center for Prevention of Preterm Birth, Perinatal Institute and March of Dimes Prematurity Research Center Ohio Collaborative, Cincinnati Children's Hospital Medical Center, Department of Pediatrics, University of Cincinnati College of Medicine, Cincinnati, Ohio, United States of America

[5]Department of Epidemiology Research, Statens Serum Institut, Copenhagen, Denmark

[6]Department of Obstetrics and Gynaecology, Sahlgrenska Academy, Institute of Clinical Science, University of Gothenburg, Gothenburg, Sweden

[7]Department of Genetics and Bioinformatics, Health Data and Digitalization, Norwegian Institute of Public Health, Oslo, Norway

[8]Institute for Molecular Medicine Finland (FIMM), Helsinki Institute of Life Science, University of Helsinki, Helsinki, Finland

[9]Program in Medical and Population Genetics, Broad Institute of Harvard and MIT, Cambridge, MA, USA.

[10]Psychiatric & Neurodevelopmental Genetics Unit, Department of Psychiatry, Analytic and Translational Genetics Unit, Department of Medicine, and the Department of Neurology, Massachusetts General Hospital, Boston, MA, USA.

[11]Center for Child, Adolescent, and Maternal Health Research, Faculty of Medicine and Health Technology, University of Tampere, Tampere, Finland

[12]Department of Obstetrics and Gynecology, Tampere University Hospital, Tampere, Finland

[13]Burroughs Wellcome Fund, Research Triangle Park, Durham, NC, USA.

[14]Faculty of Medicine and Health Technology, Tampere University, Tampere, Finland

*Corresponding author

**equal contribution

### Contents

**Supplementary Figure 1.** Meta-analysis of gestational duration and SPTB with a strict definition of spontaneous birth in the FinnGen-based GWAS.

**Supplementary Figure 2.** Effect estimates for the genome-wide significant meta-analysis loci in the FinnGen based GWAS.

**Supplementary Figure 3.** QQ-plots of the meta-analysis of gestational duration and SPTB.

**Supplementary Figure 4.** Effect estimates of the genome-wide significant loci in the meta-analysis populations of SPTB.

**Supplementary Figure 5.** Gene expression of GA-and SPTB-annotated genes across 30 tissue types in GTEx v8.

**Supplementary Figure 6.** Regional association plots of the new loci associated with gestational duration.

**Supplementary Figure 7.** Regional association plots of the new loci associated with SPTB.

**Supplementary figure 8.** Associations of the novel candidate genes from the meta-analysis of SPTB and gestational duration in the FinnGen R7 GWAS endpoint categories

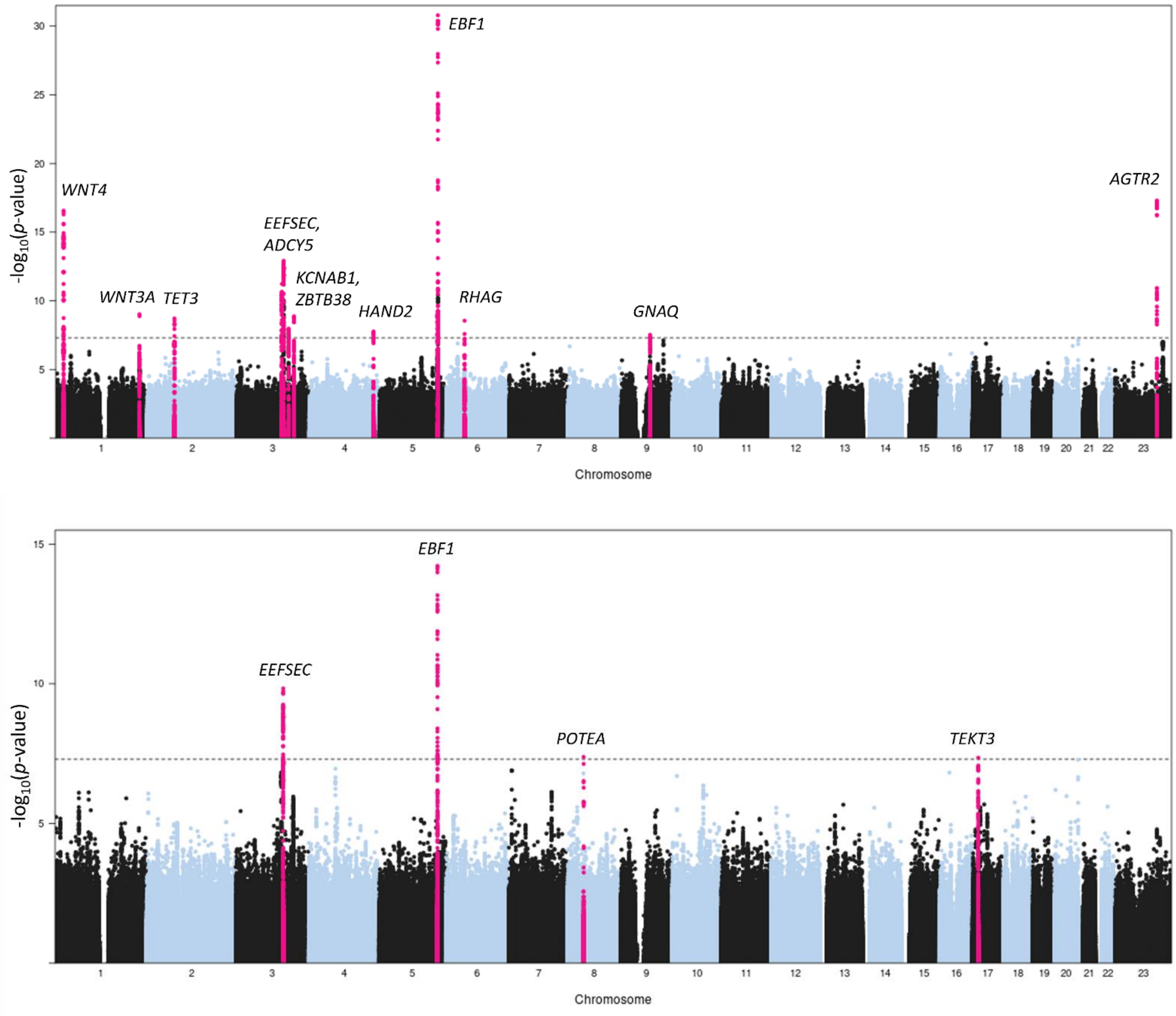


**Supplementary Figure 1**. Meta-analysis of gestational duration (upper panel) and SPTB in which a strict definition of spontaneous birth was used in the GWAS of the FinnGen data. The meta-analysis of gestational duration was performed with 66,676 samples whereas the meta-analysis of SPTB included 94,782 samples (4,953 cases and 89,829 controls).

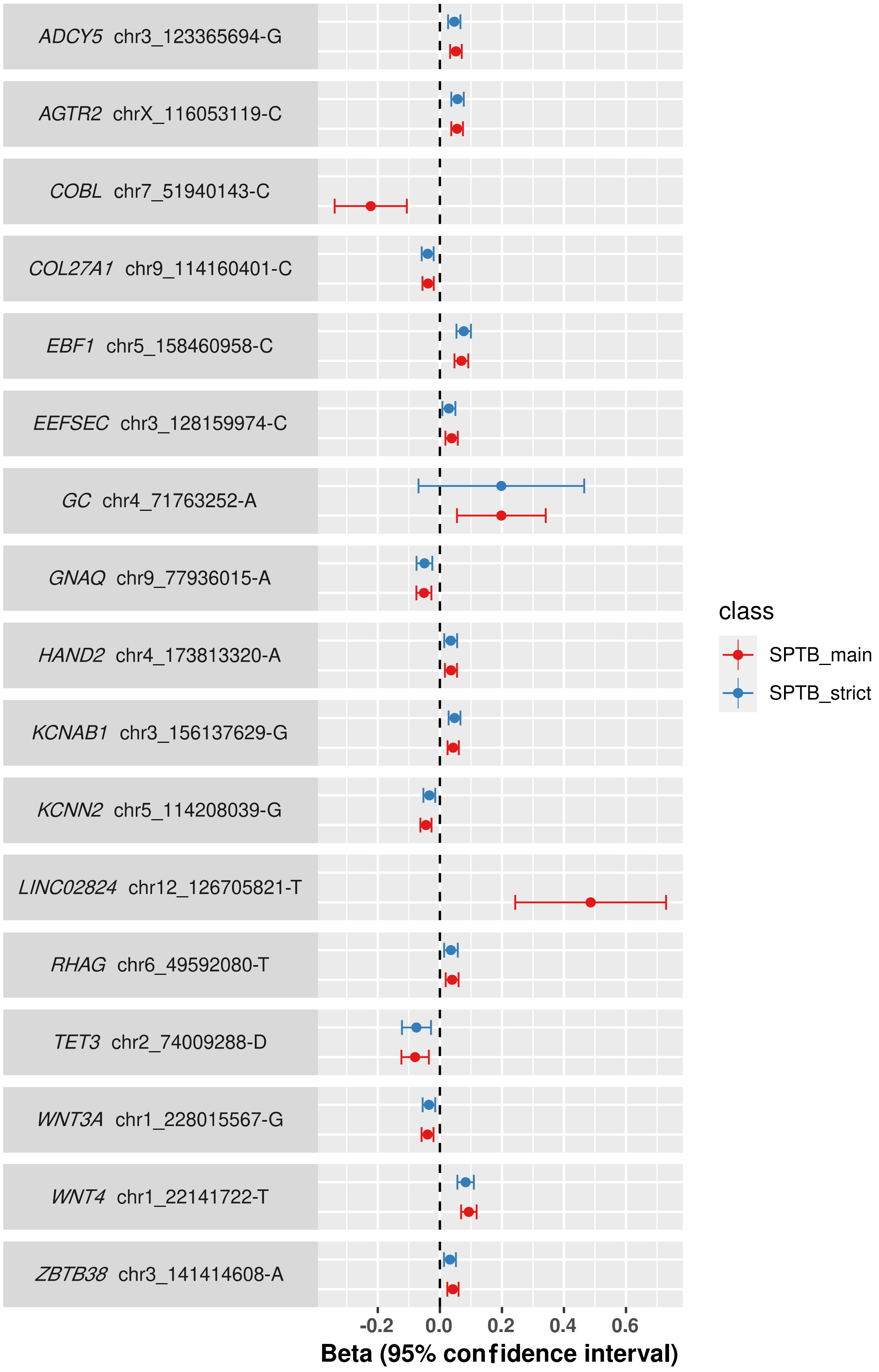


**Supplementary Figure 2.** Effect estimates for the genome-wide significant meta-analysis loci in the FinnGen based GWAS. The genes mapped to the meta-analysis risk loci for gestational duration and SPTB are shown in alphabetical order. The main GWAS of gestational duration was performed with data from 24,391 samples, and the GWAS with strict definition with 21,660 samples. The main GWAS of SPTB was based on 54,030 samples (4,925 cases and 49,105 controls), whereas the strict GWAS of SPTB had 50,441 samples (1,336 cases and 49,105 controls).

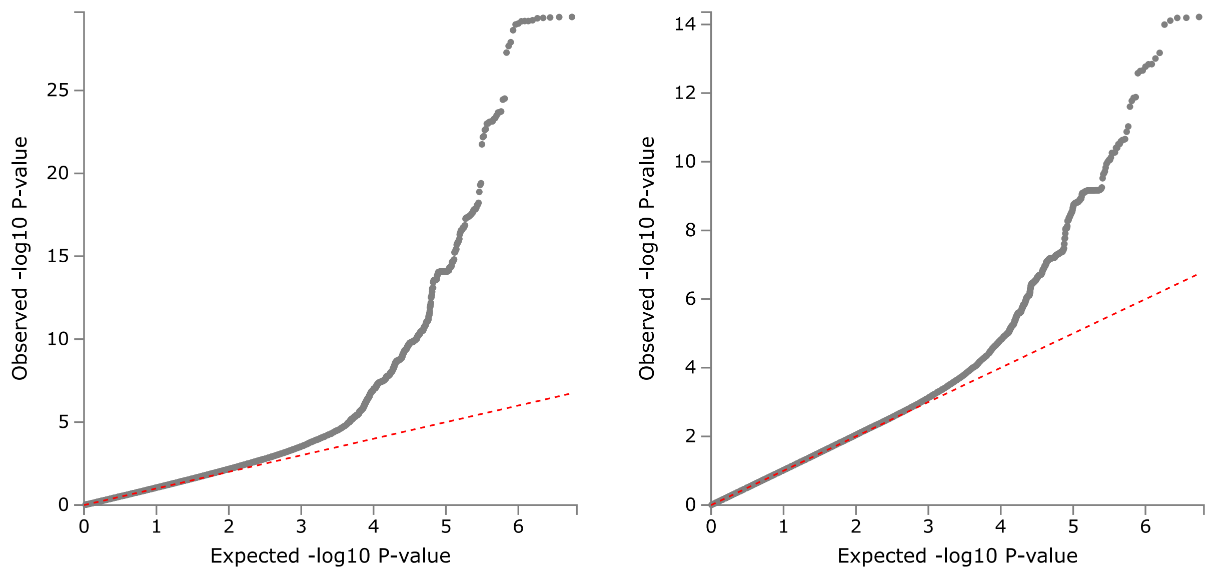


**Supplementary figure 3.** QQ-plots of the meta-analysis of gestational duration (left) and SPTB.

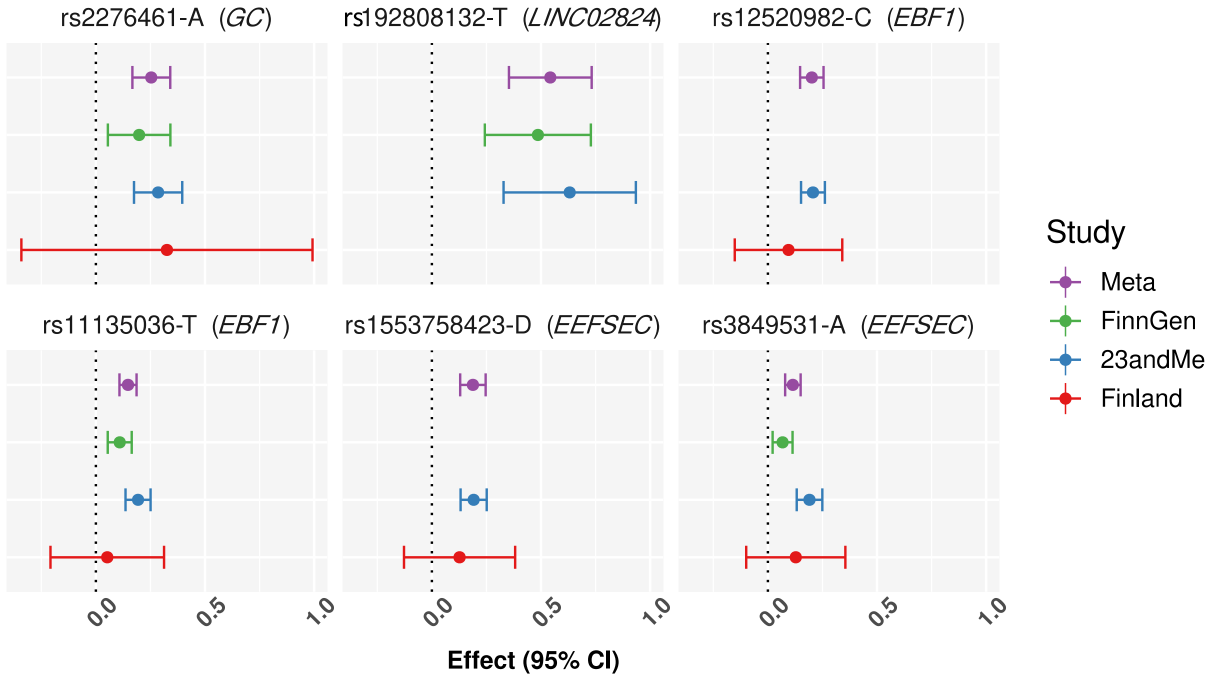


**Supplementary figure 4**. Effect estimates of the genome-wide significant loci from the meta-analysis populations of SPTB. Two variants in *EBF1* and *EEFSEC* loci are shown because the meta-analysis lead variants were not present in the FinnGen-based GWAS.

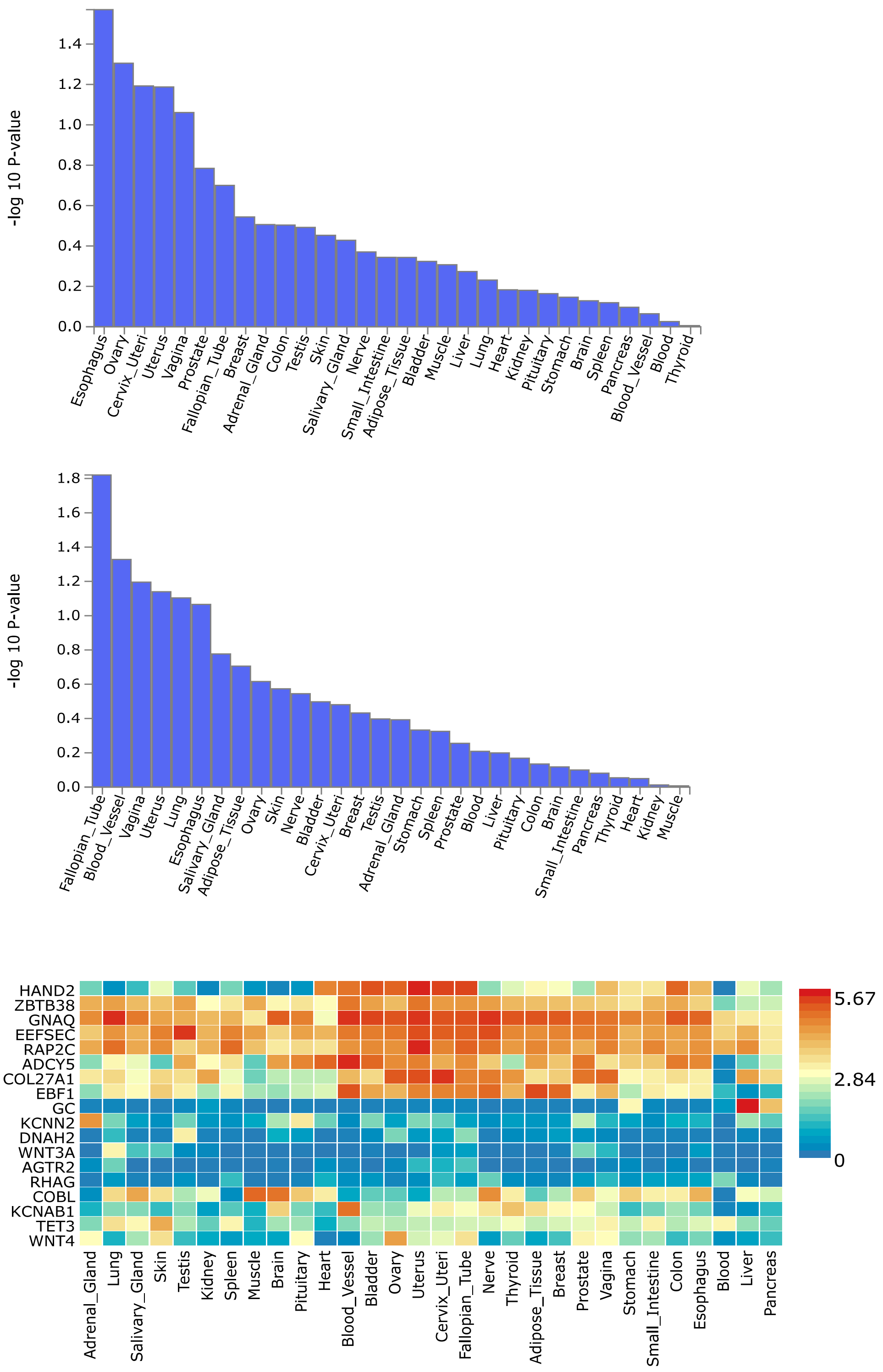


**Supplementary Figure 5**. Gene expression of gestational duration- and SPTB-annotated genes across 30 tissue types in GTEx v8. The histograms show MAGMA gene set enrichment analysis for gestational duration and SPTB, and the heatmap features gene expression depicted as averaged expression value per tissue type, with hierarchal clustering for both genes and tissues.

**Supplementary Figure 6.** Regional association plots of the novel loci associated with gestational duration.

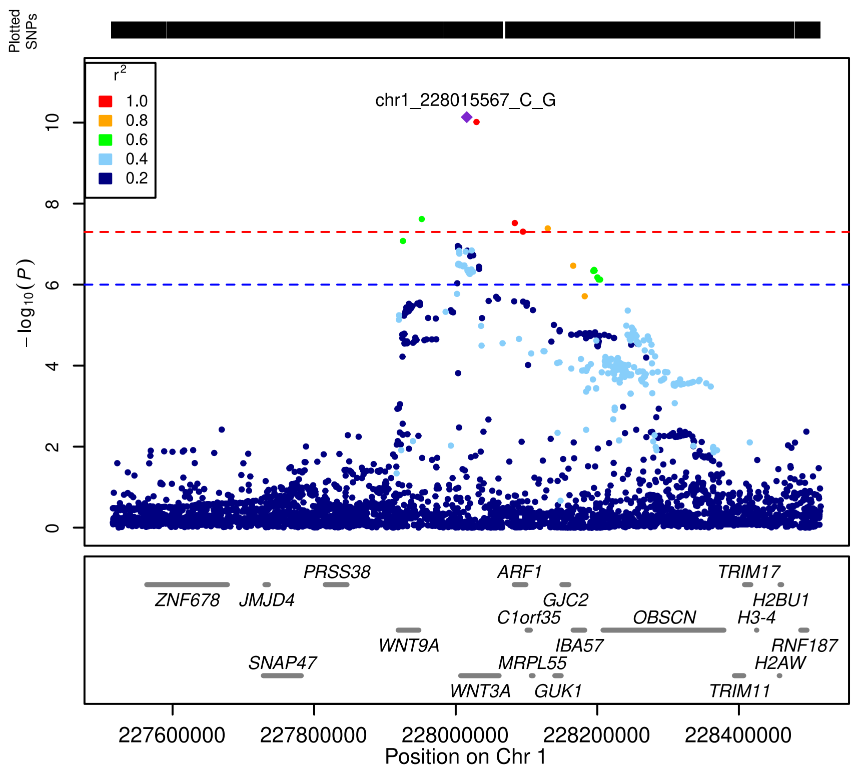


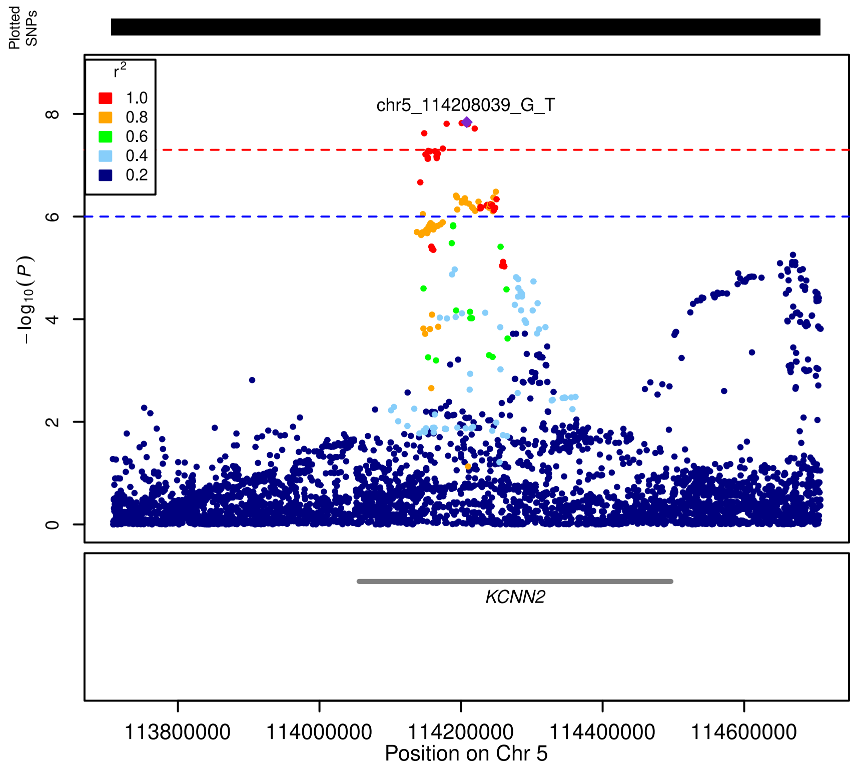

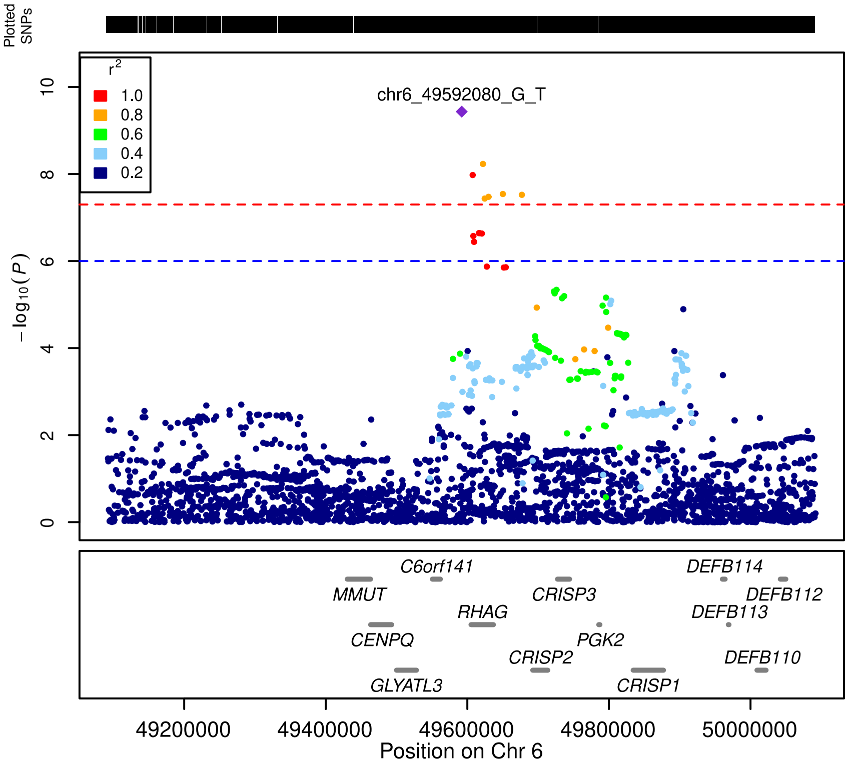
Plotted SNPs
r2
1.0
0.8
0.6
0.4
0.2
chr6_49592080_G_T
−log10(P)
0
2
4
6
8
10
C6orf141
MMUT
CRISP3
DEFB114
DEFB112
RHAG
DEFB113
CENPQ
PGK2
CRISP2
DEFB110
GLYATL3
CRISP1
49200000
49400000
49600000
49800000
50000000
Position on Chr 6

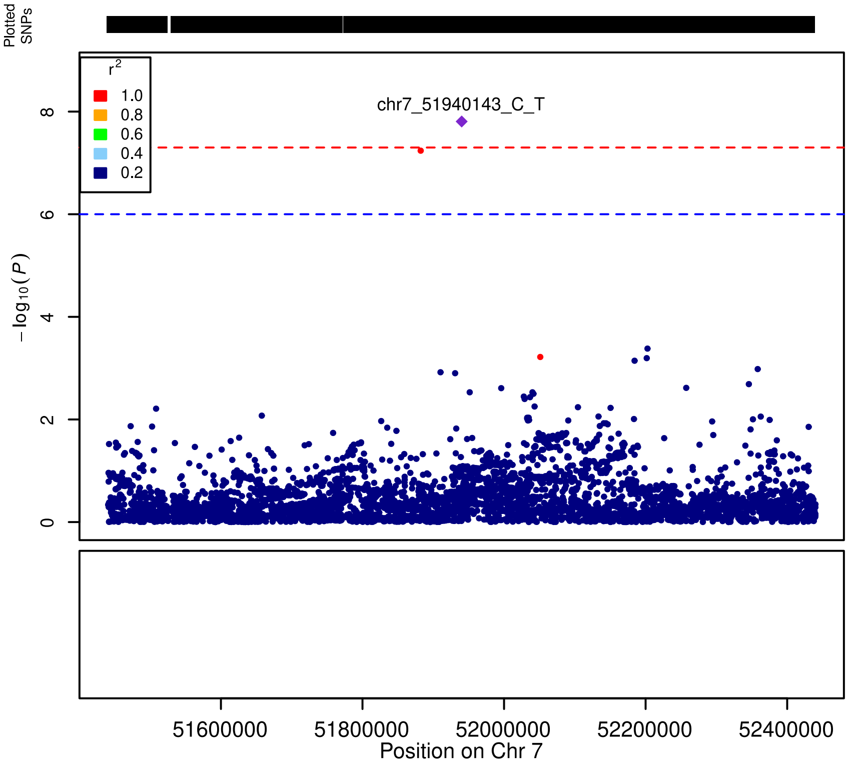
Plotted SNPs
r2
1.0
0.8
0.6
0.4
0.2
chr7_51940143_C_T
−log10(P)
0
2
4
6
8
51600000
51800000
52000000
52200000
52400000
Position on Chr 7

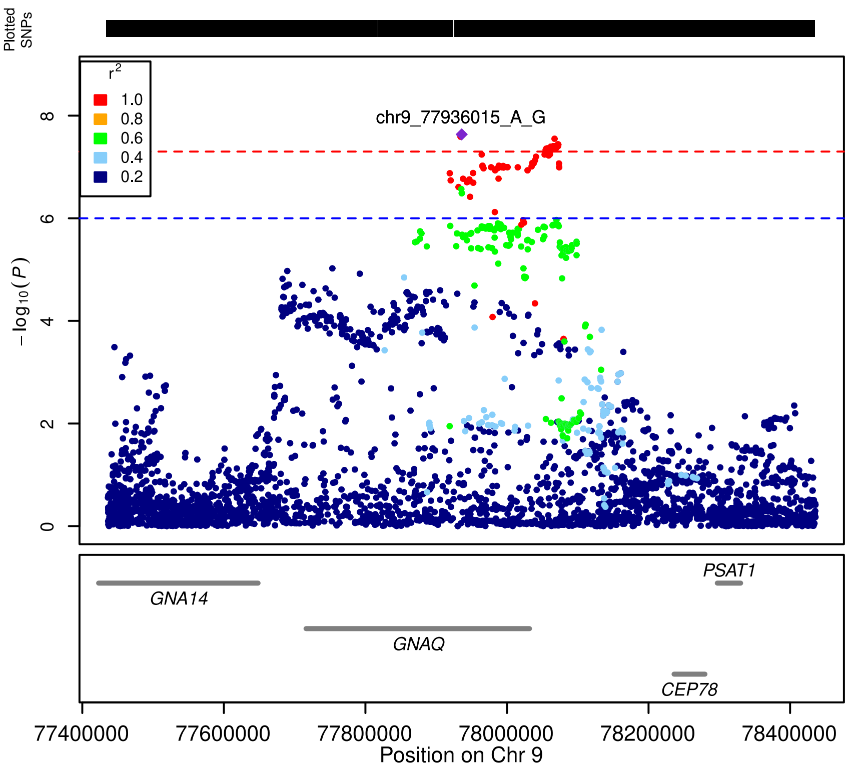

Plotted SNPs
r2
1.0
0.8
0.6
0.4
0.2
chr9_77936015_A_G
0
2
4
6
8
−log10(P)
GNA14
GNAQ
PSAT1
CEP78
77400000
77600000
77800000
78000000
78200000
78400000
Position on Chr 9

**Supplementary Figure 7.** Regional association plots of the novel loci associated with SPTB.

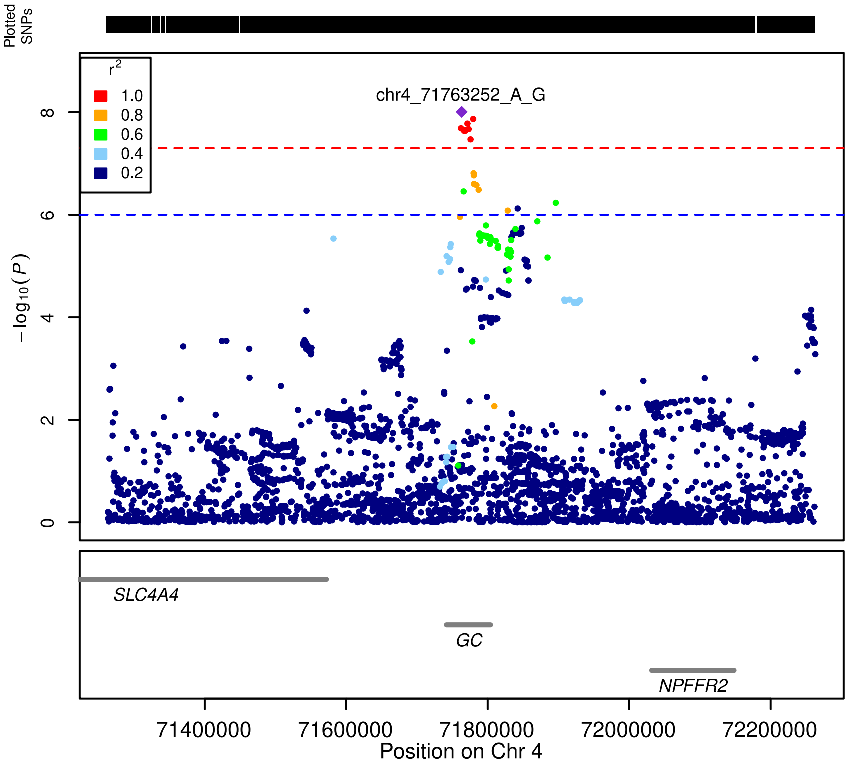


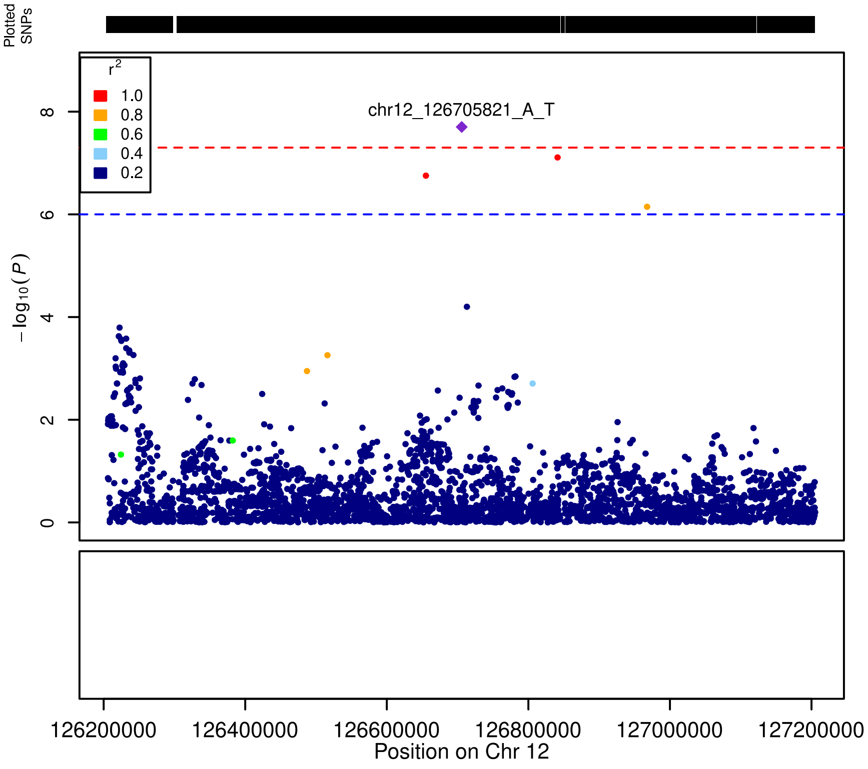

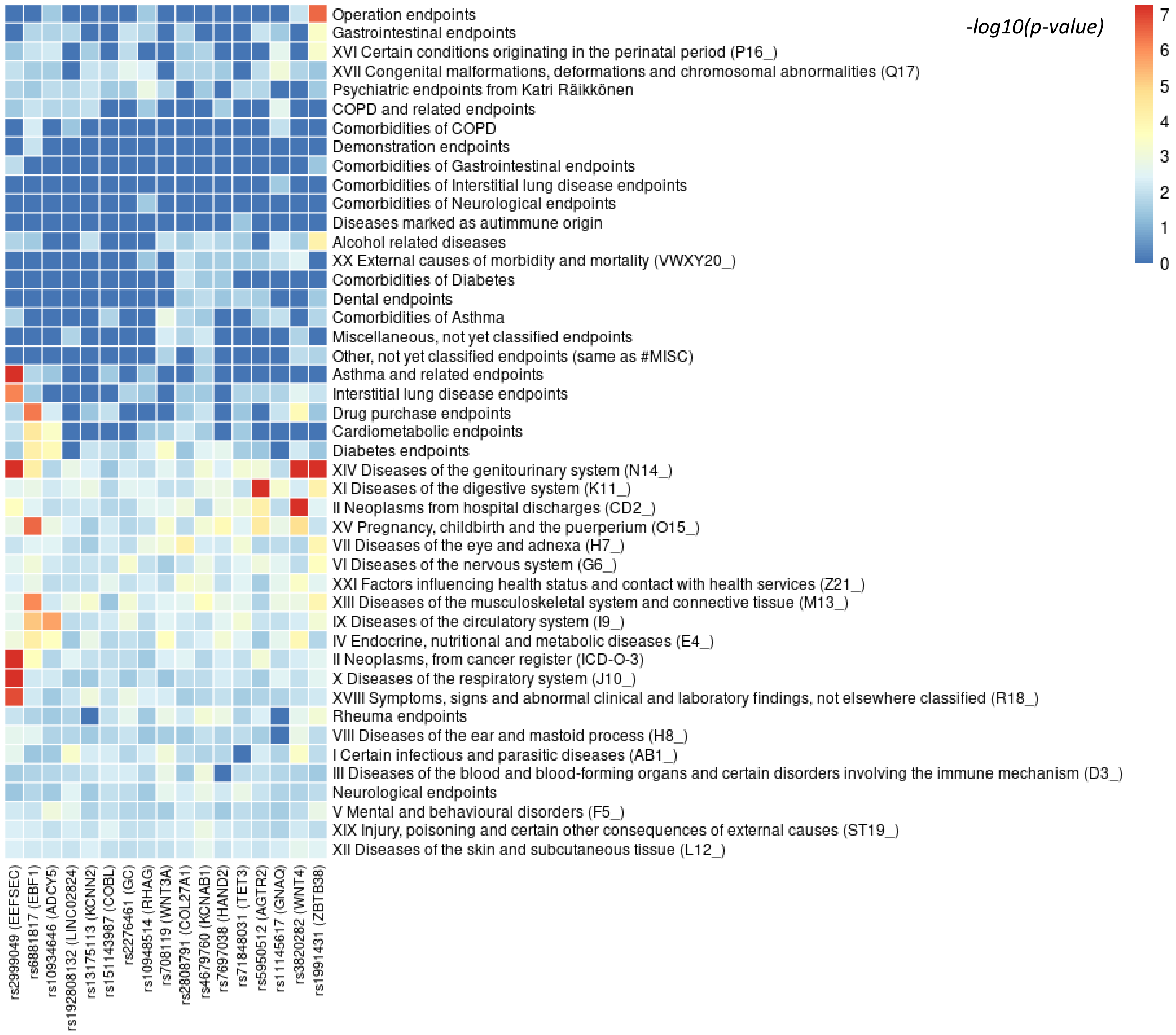


**Supplementary Figure 8**. Associations of the candidate genes from the meta-analysis of SPTB and gestational duration in the FinnGen R7 GWAS endpoint categories, each comprising >3,000 traits. In each category, *p* value is based on the strongest associating trait.
